# Change in age distribution of COVID-19 deaths with the introduction of COVID-19 vaccination

**DOI:** 10.1101/2021.07.20.21260842

**Authors:** Roberta Pastorino, Angelo Maria Pezzullo, Leonardo Villani, Francesco Andrea Causio, Cathrine Axfors, Despina G. Contopoulos-Ioannidis, Stefania Boccia, John P.A. Ioannidis

## Abstract

**Background:** Most countries initially deployed COVID-19 vaccines preferentially in elderly populations. Population-level vaccine effectiveness may be heralded by an increase in the proportion of deaths among non-elderly populations that were less covered by vaccination programs.

**Methods:** We collected data from 40 countries on age-stratified COVID-19 deaths during the vaccination period (1/14/2021-5/31/2021) and two control periods (entire pre-vaccination period and excluding the first wave). We meta-analyzed the proportion of deaths in different age groups in vaccination versus control periods in countries with low vaccination rates; (2) countries with age-independent vaccination policies; and (3) countries with standard age-dependent vaccination policies.

**Findings:** Countries that prioritized vaccination among older people saw an increasing share of deaths among 0-69 year old people in the vaccination versus the two control periods (summary prevalence ratio 1·32 [95 CI% 1·24-1·41] and 1·35 [95 CI% 1·26-1·44)]. No such change was seen on average in countries with age-independent vaccination policies (1·05 [95 CI% 0·78-1·41 and 0·97 [95 CI% 0·95-1·00], respectively) and limited vaccination (0·93 [95 CI% 0·85-1·01] and 0·95 [95 CI% 0·87-1·03], respectively). Prevalence ratios were associated with the difference of vaccination rates in elderly versus non-elderly people. No significant changes occurred in the share of deaths in age 0-49 among all 0-69 deaths in the vaccination versus pre-vaccination periods.

**Interpretation:** The substantial shift in the age distribution of COVID-19 deaths in countries that rapidly implemented vaccination predominantly among elderly may herald the population level-effectiveness of COVID-19 vaccination and a favorable evolution of the pandemic towards endemicity with fewer elderly deaths.

**Funding:** This study received no specific funding.

## INTRODUCTION

Several COVID-19 vaccines have shown very high efficacy in clinical trials (1–3) and have also demonstrated high effectiveness in preliminary analyses after wide deployment in large populations (4–6). While they are diminishing the total mortality footprint from the epidemic waves, vaccines may also be expected to shift the demographics of COVID-19 fatalities, especially if the deployment of vaccines happens at different pace and intensity in different demographic groups.

The aim of the study was to assess whether the proportion of deaths accounted by non-elderly has increased in the mid-January through late May 2021 time period as compared with the previous phase of the pandemic in countries that have proceeded with vaccine roll-out in 2021 and that have detailed information of the age distribution of fatalities in different periods. The rationale was that almost all countries had a vaccination roll-out policy that started from the elderly and nursing homes (7, 8). In most countries, very few young and middle age people were vaccinated in the first three months of roll out. Therefore, in the large majority of countries that used vaccines, the rates of vaccination were much higher in elderly versus non-elderly in early 2021. However, the exact difference in vaccination rates between these two age strata varied a lot, mostly because of different rapidity in deploying vaccines. If vaccines are very effective, one expects that the proportion of COVID-19 deaths accounted by elderly has decreased (and, conversely, the proportion of COVID-19 deaths accounted by non-elderly has increased) in the first 5 months of 2021; and that the change may depend on the differential rate of vaccination between elderly and non-elderly strata. Based on the same rationale, it was expected that countries that had no substantial roll-out of vaccines in the same period in any age group and those that aimed to vaccinate from the very early days all vulnerable people regardless of age would not experience a shift in the age distribution of COVID-19 deaths. Evaluation of demographic change patterns would therefore offer an indirect way of assessing the effectiveness of vaccination at the population level. We set out to assess these patterns using data from 40 countries with >500 COVID-19 deaths by end of May 2021.

## METHODS

### Data on COVID-19 deaths and definitions of contrasted time periods

We considered all countries that had a total of COVID-19 deaths > 500 as of end of May 2021, and which had publicly-available age-stratified information on COVID-19 deaths so as to separate elderly versus non-elderly age strata with a cut-off at 70 years (or 65 or 60, if 70 years cut-off data were unavailable). The selected cut-off is similar to what was used in a previous analysis of age distribution in the first versus second wave of the pandemic until January 2021 (9).

We collected data on deaths from Institut national d’études démographiques (INED) (9), COVerAGE-DataBase (10), and other national databases (11–14). England and Wales were grouped because their mortality data were reported together. All the sources about deaths are shown in supplementary table S1. Of 122 countries with >500 COVID-19 deaths (15) eligible age-stratified data were eventually available for 40 countries.

We calculated the number of deaths from January 14, 2021 (or as close as possible if data on this date were unavailable) until the latest date with data available at the time of our searches (between the end of May and the beginning of June 2021); this is called the “vaccination period”. January 14, 2021 was selected, because, while vaccinations started in December 2020 in most of the analyzed countries, any meaningful benefit is unlikely to have been seen at the population level by early January. For comparison, we used two control periods preceding the vaccination period: the “entire pre-vaccination period”, i.e. from the onset of the pandemic; and the “pre-vaccination second wave period” which only considered deaths during the second wave of the pandemic up until the start of the vaccination period. Deaths during the latter control period offer a more proximal comparison to vaccination period and they were calculated by excluding deaths that had occurred in the first wave. The first wave period was defined as in a previous analysis (16) and the data on deaths during the first wave were taken directly from that analysis and updated if more recently data were available. For the countries that had not been included in that previous analysis, we used for consistency as the separating date between the first and second waves the date with trough (lowest number) of deaths for a 7-day average according to Worldometer (15). When data were not available specifically up to the trough date, we considered a separation date for the two waves that was as close as possible to the trough. Information about the trough and the separation dates are shown in table S1.

Age-stratified death counts for the time periods of interest were separately recorded for three age groups: 0-49, 50-69, and ≥ 70 years. Data were instead available for the age groups 0-44, 45-64, ≥ 65 for Austria, Belgium, and USA; 0-39, 40-59, ≥ 60 for Bangladesh and Northern Ireland; and 0-44, 45-59 and ≥ 60 for Indonesia. Deaths in chronic care establishments and in other social or medico-social residential facilities in France were counted in the ≥ 70 age group.

There were no data available on deaths that occurred in the second wave pre-vaccination period with suitable age stratification for Canada, Israel, Jamaica, Jordan, Northern Ireland and Uruguay.

### Vaccination data

We obtained data on COVID-19 vaccination coverage (at least one dose received and full vaccination) from international databases for the general population and for the 0-69 and ≥ 70 year age groups. Number of vaccinations in the age group was extracted from the cited databases, and if the corresponding population denominator was not provided by the source, we derived it from publicly available population pyramids (17). Sources and vaccination coverage by age groups 0-69, ≥ 70 and overall are available in the supplementary table S2. In order to consider the effect of COVID-19 vaccination on deaths, we used the vaccination coverage until April 30, 2021 (or as close as possible if data on this date were unavailable). This date was chosen to allow a 1-month difference between the vaccination data and the end of the period during which deaths were counted.

### Contrasts of age groups and data synthesis

The main analysis compared the proportion of COVID-19 deaths in the age group 0-69 (0-59 or 0-64 when these were not available)/per total COVID-19 deaths (prevalence) in the vaccination period versus each of the two pre-vaccination control periods. Prevalence ratios for each country (proportion of COVID-19 deaths accounted by 0-69 year old people in the vaccination period divided by the proportion of COVID-19 deaths accounted by the same age stratum in a control period for the same country) were calculated along with 95% confidence intervals and were synthesized across countries with inverse-variance random effects meta-analysis. Between-country heterogeneity was measured using the I^2^ statistic and tested with the Q test. The meta-analysis was also stratified in three categories: 1) countries with low vaccination rates, i.e. proportion of vaccination with at least one dose < 5% in the general population by late April 2021 (Japan, Republic of Moldova, Peru, Ukraine, Philippines, Bangladesh, Jamaica, and Indonesia); 2) countries with age-independent policies, i.e. adopting very early (before 1/15/2021) vaccination policies for vulnerable groups regardless of age (Romania, Denmark, Israel) (18); 3) countries with standard age-dependent policies (Argentina, Austria, Belgium, Brazil, Canada, Colombia, England and Wales, Finland, France, Germany, Greece, Hungary, Italy, Jordan, Mexico, Netherlands, Northern Ireland, Norway, Poland, Portugal, Scotland, Slovenia, South Korea, Spain, Sweden, Switzerland, Uruguay, and USA). As discussed above, a change in the age demographics of deaths was expected to have occurred only in the third group. Since Australia registered only one death in the vaccination period it was excluded from analyses.

A secondary analysis compared the proportion of COVID-19 deaths in the age group 0-49 years (0-39 or 0-44 when these were not available) among all COVID-19 deaths in the age group 0-69 (0-59 or 0-64 when these were not available) in the vaccination period versus each of the two pre-vaccination control periods. Vaccination rates were limited in people < 70 years old by April 2021 and therefore, no change in the age demographics was expected to have occurred when these periods were compared. Meta-analysis of prevalence ratios across countries used random effects similar to the primary analysis.

Finally,we performed exploratory meta-regression analyses, where we examined if the natural logarithm of the prevalence ratio of deaths in the 0-69 age group in the vaccination versus each control period was associated with the difference in the proportion of people who were vaccinated by the end of April in those ≥ 70 versus those 0-69 years old (using a 60 or 65 years cut-off, if data for the 70 years cut-off were not available). This difference shows the strength of the contrast in vaccination rates between the two age groups of interest and we hypothesized that it may explain in part the differential change in deaths in the two age strata. The meta-regression analysis was weighted by the inverse of the variance of the natural logarithm of the prevalence ratio. We considered two different versions of the meta-regression, one using the data on the proportion of people who were fully vaccinated and another using the data on the proportion of people who had received at least one dose, given that vaccine efficacy may be substantial even within 1-2 weeks after a single dose (5, 6). For the countries of Group 1 (those with low vaccination rates) we had vaccinations per age strata only for Japan and Peru, while for the countries of Group 3 we had missing data for six countries. We imputed the missing values in each group as the median of the differences in the proportion of vaccinated people between the relevant age strata of the countries with observed data in their group. The countries of Group 2 that very early vaccinated all vulnerable groups regardless of age (Romania, Denmark, and Israel) were excluded from this analysis.

### Statistical analyses

All analyses were performed in STATA 16·0 (StataCorp, 2019). We used P<0·005 as the threshold for statistical significance and values between 0·005 and 0·05 were considered suggestive (19).

## RESULTS

### Deaths and vaccination rates per age strata

Table 1 reports the number of COVID-19 deaths in people 0-69 years old and the total COVID-19 deaths in each of the 40 eligible countries with data for the two control periods, and the vaccination period. The proportion of COVID-19 deaths that had occurred in people 0-69 years old ranged from 5·0% to 67·1% in the entire pre-vaccination period, from 5·2% to 67·2% in the second wave pre-vaccination period, and from 0·0% to 64·4% in the vaccination period (median 15·6%, 14·7%, and 20·6%, respectively). The proportions were much lower in high-income countries (range 5·0-30·5% [median 11·8%], 5·2-30·6% [median 11·5%], and 0·0%-36·0% [median 15·3%], in the three periods, respectively).

**Table 1.** Proportions of COVID-19 deaths in the 0-69 age group to total COVID-19 deaths (%), separately for entire pre-vaccination period, second wave pre-vaccination period, and vaccination period.

| Country | Entire pre-vaccination period | Second wave pre-vaccination period | Vaccination period |
| --- | --- | --- | --- |
| Argentina | 17,597/47,665 (36.9) | 8877/23,524 (37.7) | 14,732/31,625 (46.6) |
| Australia | 58/909 (6.4) | 44/812 (5.4) | 0/1 (0.0) |
| Austria | 422/6828 (6.2) | 384/6183 (6.2) | 374/3485 (10.7) |
| Bangladesh | 4781/7835 (61.0) | 4309/7086 (60.8) | 2172/3560 (61.0) |
| Belgium | 1232/20,224 (6.1) | 694/10,613 (6.5) | 442/4418 (10.0) |
| Brazil | 94,070/212,007 (44.4) | 21,887/50,274 (43.5) | 151,324/256,026 (59.1) |
| Canada* | 1883/17,315 (10.9) | - | 1593/7692 (20.7) |
| Colombia | 24,207/55,034 (44.0) | 9570/23,420 (40.9) | 24,432/47,669 (51.3) |
| Denmark | 203/1720 (11.8) | 128/1122 (11.4) | 88/791 (11.1) |
| England and Wales | 15,104/94,132 (16.0) | 6722/42,493 (15.8) | 9037/44,033 (20.5) |
| Finland | 67/594 (11.3) | 33/286 (11.5) | 72/390 (18.5) |
| France | 7397/69,044 (10.7) | 3702/38,816 (9.5) | 5684/39,706 (14.3) |
| Germany | 4629/41,486 (11.2) | 3303/32,343 (10.2) | 6734/45,749 (14.7) |
| Greece | 835/5421 (15.4) | 793/5246 (15.1) | 1091/6495 (16.8) |
| Hungary | 3047/11,066 (27.5) | 2918/10,477 (27.9) | 6635/18,708 (35.5) |
| Indonesia | 14,137/25,245 (56.0) | 10,382/19,173 (54.1) | 11,819/26,199 (45.1) |
| Israel* | 770/3704 (20.8) | - | 728/2630 (27.7) |
| Italy | 10,974/78,591 (14.0) | 5759/42,947 (13.4) | 7178/44,684 (16.1) |
| Jamaica* | 141/312 (45.2) | - | 312/638 (48.9) |
| Japan | 421/3338 (12.6) | 294/2512 (11.7) | 657/6428 (10.2) |
| Jordan* | 1406/4040 (34.8) | - | 2097/5469 (38.3) |
| Mexico | 102,687/156,933 (65.4) | 39,175/62,379 (62.8) | 37,297/60,412 (61.7) |
| Netherlands | 1215/12,563 (9.7) | 520/6428 (8.1) | 658/5003 (13.2) |
| Northern Ireland* | 76/1532 (5.0) | - | 39/621 (6.3) |
| Norway | 76/511 (14.9) | 47/276 (17.0) | 71/272 (26.1) |
| Peru | 52,794/96,610 (54.6) | 3233/6477 (49.9) | 50,240/87,885 (57.2) |
| Philippines | 7724/11,509 (67.1) | 6864/10,214 (67.2) | 5625/8731 (64.4) |
| Poland | 8036/26,371 (30.5) | 7575/24,775 (30.6) | 12664/39,260 (32.3) |
| Portugal | 1050/8543 (12.3) | 811/6793 (11.9) | 1158/8479 (13.7) |
| Republic of Moldova | 1743/2942 (59.2) | 1190/2058 (57.8) | 1418/2663 (53.2) |
| Romania | 7251/17,021 (42.6) | 6268/15,739 (39.8) | 5077/13,096 (38.8) |
| Scotland | 727/7467 (9.7) | 346/3237 (10.7) | 405/2647 (15.3) |
| Slovenia | 177/3423 (5.2) | 172/3313 (5.2) | 112/1270 (8.8) |
| South Korea | 189/1195 (15.8) | 138/970 (14.2) | 132/745 (17.7) |
| Spain | 7831/57,080 (13.7) | 3638/27,376 (13.3) | 3800/22,417 (17.0) |
| Sweden | 878/9940 (8.8) | 291/4248 (6.9) | 510/3642 (14.0) |
| Switzerland | 638/7904 (8.1) | 473/6264 (7.6) | 285/2346 (12.1) |
| Ukraine | 10,382/20,305 (51.1) | 9916/19,524 (50.8) | 12,230/26,849 (45.6) |
| Uruguay* | 77/280 (27.5) | - | 1353/3755 (36.0) |
| USA | 79,806/416,640 (19.2) | 53,136/285,179 (18.6) | 37,153/162,388 (22.9) |

\* Deaths that occurred in the second wave were not available in suitable age stratification.

Table 2 reports the data about vaccination coverage in each of the 40 countries by the end of April in the 0-69 and ≥ 70 years old age strata and in the overall population. The proportion of the general population that had been fully vaccinated varied from 0·0% to 57·4% across the 40 countries, while for those who had received at least one dose the proportion varied from 1·5% to 61·4% across the 40 countries. The proportion of the population 0-69 years old that had been fully vaccinated ranged from 1·3% to 53·8%, and 1·5% to 57·9% had received at least one dose. Conversely, the proportion of the population >=70 years old with full vaccination ranged from 0·0% to 99·7%, and 0·6%-100·0% had received at least one dose.

**Table 2.** COVID-19 vaccination coverage by <u>age groups and overal</u>l (%).

| Country | 0-69 |  | ≥ 70 |  | Overall |  | Date |
| --- | --- | --- | --- | --- | --- | --- | --- |
|  | At least one dose | fully vaccinated | At least one dose | fully vaccinated | At least one dose | fully vaccinated |  |
| Argentina | 10.3 | 1.8 | 100.0<sup>a</sup> | 7.9 | 15.9 | 2.2 | Apr 30 2021 |
| Australia | NA | NA | NA | NA | NA | NA | Apr 30 2021 |
| Austria | 20.0 | 5.9 | 69.8 | 35.7 | 23.5 | 9.0 | Apr 25 2021 |
| Bangladesh | NA | NA | NA | NA | 3.5 | 1.7 | Apr 29 2021 |
| Belgium | 13.9 | 5.0 | 86.4 | 17.6 | 23.9 | 6.7 | Apr 25 2021 |
| Brazil | 8.9 | 3.2 | 98.3 | 69.3 | 14.1 | 7.0 | Apr 30 2021 |
| Canada | 21.2 | 1.6 | 85.6 | 8.7 | 29.3 | 2.5 | Apr 24 2021 |
| Colombia | NA | NA | NA | NA | 6.6 | 3.0 | Apr 30 2021 |
| Denmark | 9.6 | 4.9 | 89.4 | 40.5 | 21.1 | 10.0 | Apr 25 2021 |
| England | 42.6 | 9.8 | 100.0<sup>a</sup> | 82.3 | 50.0 | 19.5 | Apr 25 2021 |
| Finland | 16.0 | 2.1 | 91.6 | 6.7 | 28.0 | 2.9 | Apr 25 2021 |
| France | 12.7 | 2.9 | 70.6 | 39.2 | 21.2 | 8.2 | Apr 25 2021 |
| Germany | NA | NA | NA | NA | 23.7 | 7.2 | Apr 25 2021 |
| Greece | 12.6 | 3.2 | 61.9 | 33.3 | 18.7 | 7.6 | Apr 25 2021 |
| Hungary | 31.0 | 11.4 | 75.4 | 53.8 | 36.9 | 17.0 | Apr 25 2021 |
| Indonesia | NA | NA | NA | NA | 4.3 | 2.5 | Apr 25 2021 |
| Israel | 57.9 | 53.8 | 100.0<sup>a</sup> | 99.7 | 61.4 | 57.4 | Apr 30 2021 |
| Italy | 10.7 | 4.5 | 64.7 | 28.5 | 20.9 | 8.7 | Apr 25 2021 |
| Jamaica | NA | NA | NA | NA | 4.9 | 0.2 | May 14 2021 |
| Japan<sup>o</sup> | 2.1 | 1.5 | 0.6 | 0.0 | 1.8 | 1.2 | Apr 30 2021 |
| Jordan | NA | NA | NA | NA | 6.9 | 2.2 | Apr 29 2021 |
| Mexico* | 1.5 | 1.3 | 73.6 | 38.4 | 9.6 | 5.5 | Apr 30 2021 |
| Netherlands | NA | NA | NA | NA | 25.6 | 6.9 | Apr 25 2021 |
| Northern Ireland | NA | NA | NA | NA | 49.8 | 22.3 | Apr 30 2021 |
| Norway | 19.9 | 6.5 | 87.7 | 33.6 | 22.2 | 5.6 | Apr 25 2021 |
| Peru | 2.2 | 1.8 | 19.9 | 2.8 | 3.2 | 1.9 | Apr 30 2021 |
| Philippines | NA | NA | NA | NA | 1.5 | 0.3 | Apr 30 2021 |
| Poland | 14.9 | 2.9 | 62.6 | 36.1 | 20.5 | 6.8 | Apr 25 2021 |
| Portugal | 11.0 | 3.0 | 77.4 | 34.4 | 21.7 | 8.1 | Apr 25 2021 |
| Republic of Moldova | NA | NA | NA | NA | 3.2 | 0.4 | Apr 30 2021 |
| Romania* | 10.8 | 5.8 | 24.6 | 16.6 | 15.8 | 9.3 | Apr 25 2021 |
| Scotland | 43.6 | 13.2 | 100.0<sup>a</sup> | 89.2 | 51.3 | 23.6 | Apr 30 2021 |
| Slovenia | 12.9 | 2.3 | 61.5 | 46.5 | 19.6 | 8.4 | Apr 25 2021 |
| South Korea | NA | NA | NA | NA | 6.5 | 0.5 | Apr 30 2021 |
| Spain | 38.3 | 14.5 | 79.5 | 34.4 | 47.1 | 26.2 | Apr 25 2021 |
| Sweden | 31.2 | 12.4 | 90.0 | 33.0 | 40.5 | 21.7 | Apr 25 2021 |
| Switzerland | 9.9 | 4.1 | 69.2 | 45.6 | 18.4 | 10.1 | Apr 25 2021 |
| Ukraine | NA | NA | NA | NA | 1.7 | 0.0 | Apr 30 2021 |
| Uruguay<sup>o</sup> | 27.3 | 18.0 | 71.6 | 30.1 | 34.0 | 19.8 | Apr 30 2021 |
| USA<sup>o</sup> | 35.1 | 22.4 | 82.6 | 69.1 | 42.9 | 30.2 | Apr 30 2021 |

\* Age group 0-59 and ≥ 60
<sup>o</sup> Age group 0-64 and ≥ 65
<sup>a</sup> More vaccinated people than the estimated population in this age category
NA: Data were not available

### Change in age distribution over time

As shown in Figure 1, the proportion of deaths accounted by those 0-69 years old increased in the vaccination period compared with the entire pre-vaccination period (panel A) or the second wave pre-vaccination period (panel B) with summary prevalence ratios being 1·20 (95% CI, 1·14-1·27) and 1·22 (95% CI, 1·15-1·30), respectively (p<0·0001 for both) when all countries were considered, but there was extreme between-country heterogeneity. An increase of the proportion of deaths accounted by the non-elderly was seen in all countries with substantial vaccination and not early age-inclusive policies (except for Mexico) with summary prevalence ratios of 1·32 (95% CI, 1·24-1·41) and 1·35 (95% CI, 1·26-1·44), respectively, but there was still large between-country heterogeneity in the exact magnitude of the prevalence ratio. For the countries of Group 1 (Japan, Republic of Moldova, Peru, Ukraine, Philippines, Bangladesh, Jamaica, and Indonesia) where very little vaccination happened in any age group, there was no difference in the proportion of deaths accounted by 0-69 years old people on average (summary prevalence ratio 0·93 (95% CI, 0·85-1·01) and 0·95 (95% CI, 0·87-1·03), respectively). Same pattern of no overall age shift was documented for the countries of Group 2 (Denmark, Romania and Israel) (summary prevalence ratio 1·05 (95% CI, 0·78-1·41) and 0·97 (95% CI, 0·95-1·00), respectively). Between-country heterogeneity existed also in the results of countries from these two groups.

**Figure 1.**
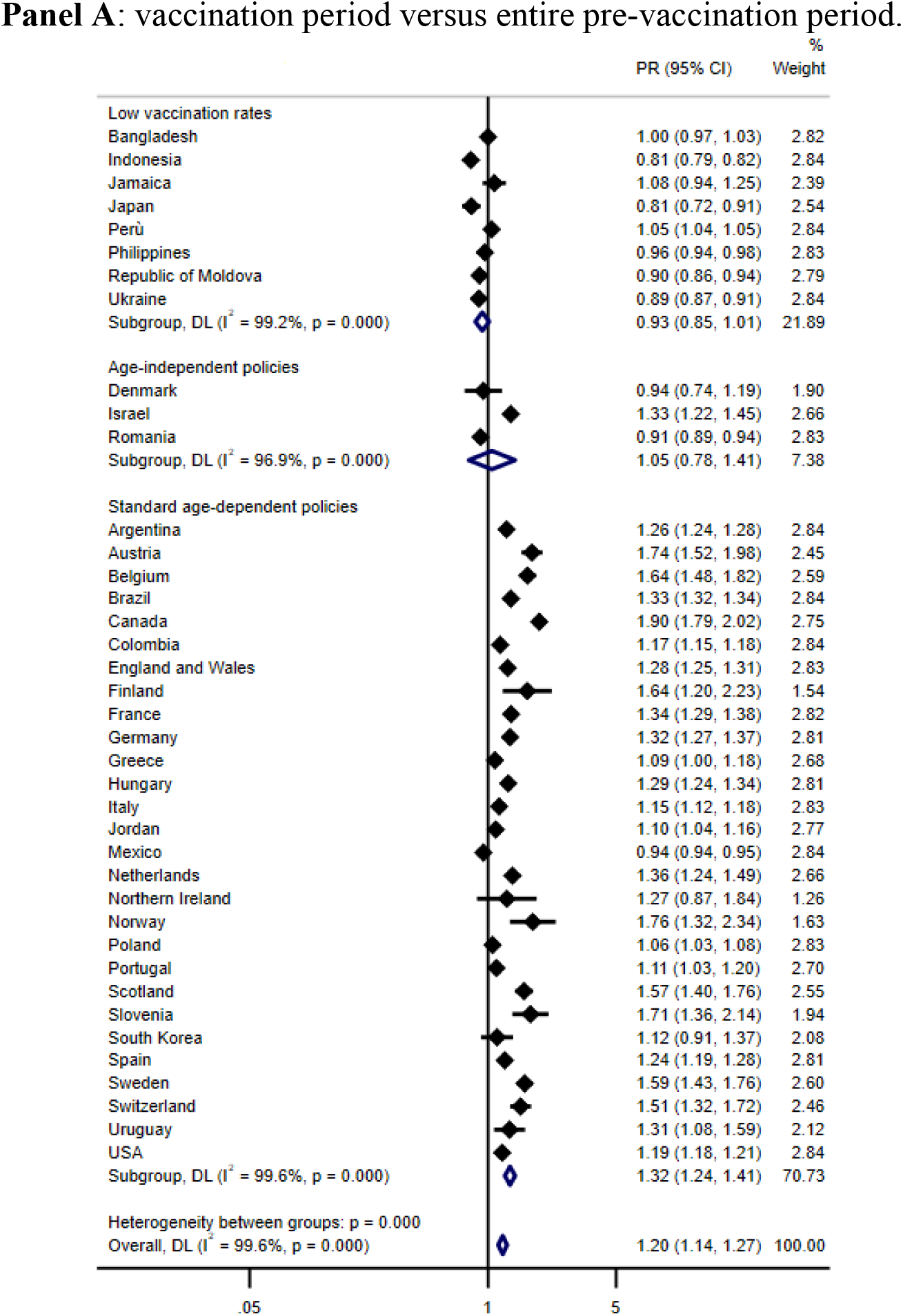
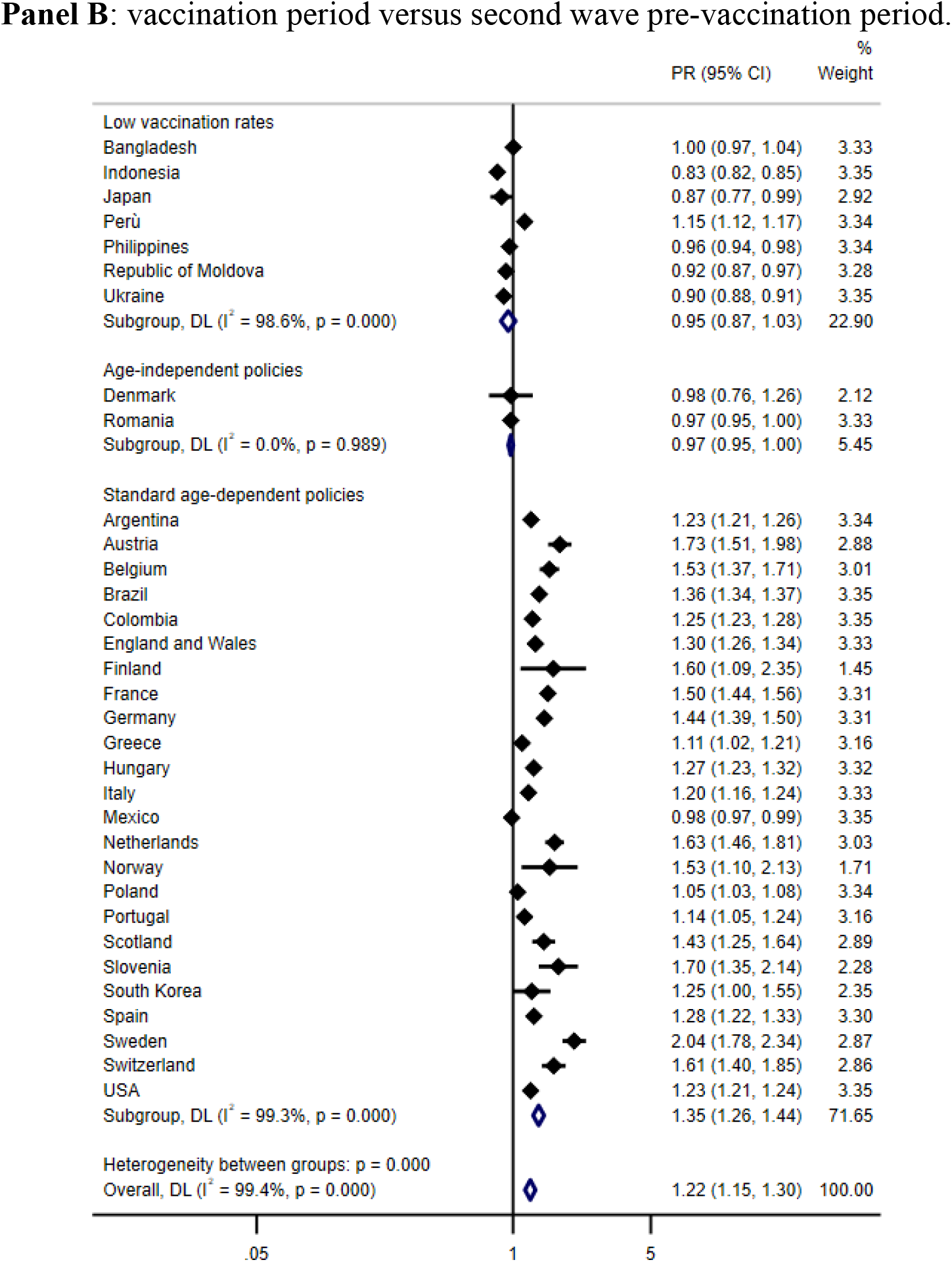
Meta-analysis of prevalence ratios (PR) of COVID-19 deaths in age 0-69 in the vaccination versus pre-vaccination control periods stratified according to three main categories 1) Low vaccination rates: countries with a proportion of vaccination with at least one dose < 5% in the general population by the end of April 2021; 2) Age-independent policies: countries with early vaccination policies that vaccinated vulnerable groups regardless of age on 15 January 2021; 3) Standard age-dependent policies: remaining countries. PR >1 implies a higher proportion of deaths accounted by 0-69 age group in the vaccination period versus pre-vaccination periods.

As shown in Figure 2, there was no significant, consistent pattern of change in the age distribution of deaths in the 0-49 age strata in the vaccination period versus the pre-vaccination control periods (summary prevalence ratio 1·00 (95% CI 0·93-1·07) and 0·99 (95% CI 0·90-1·09), p>0·05 for both).

**Figure 2.**
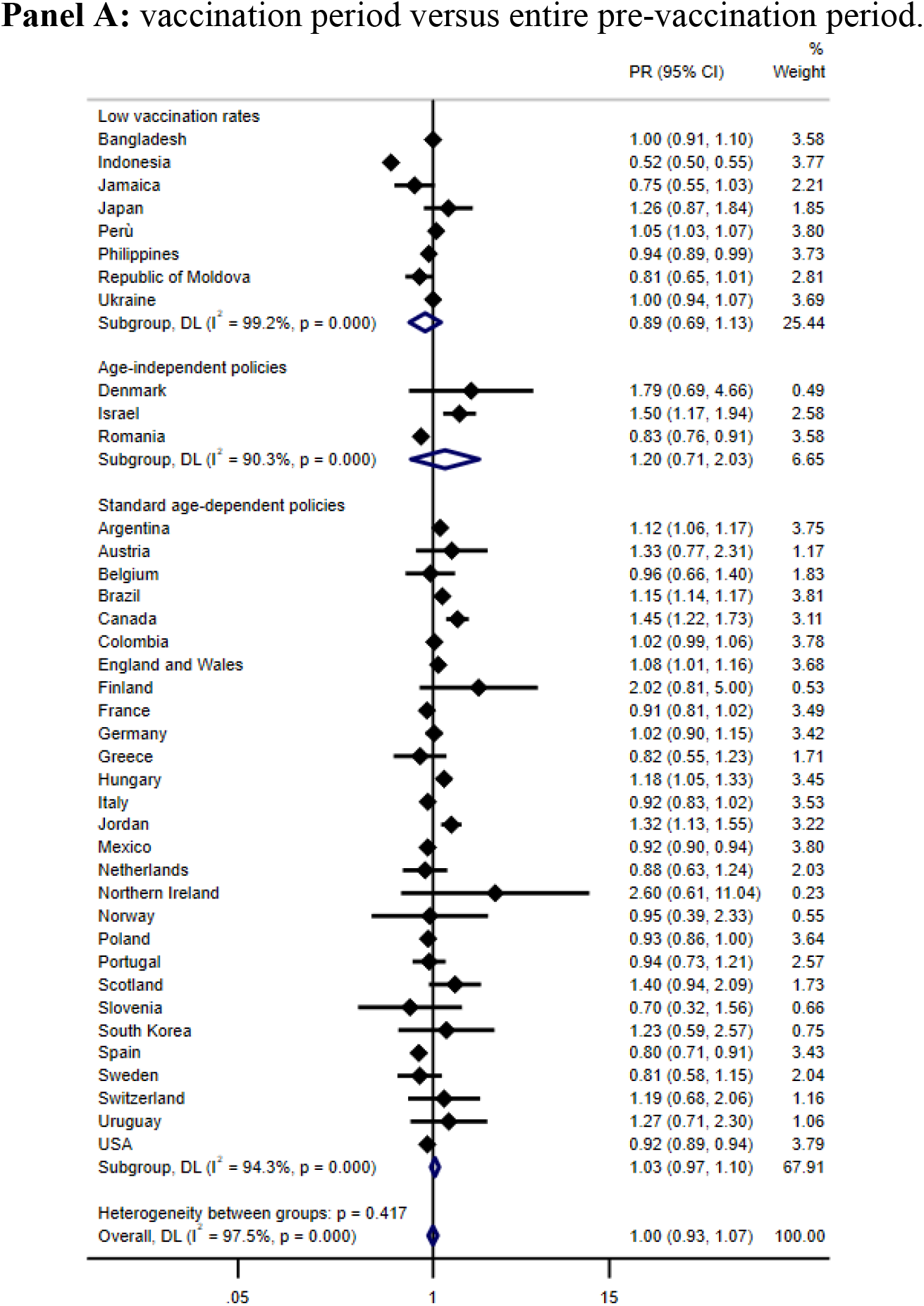
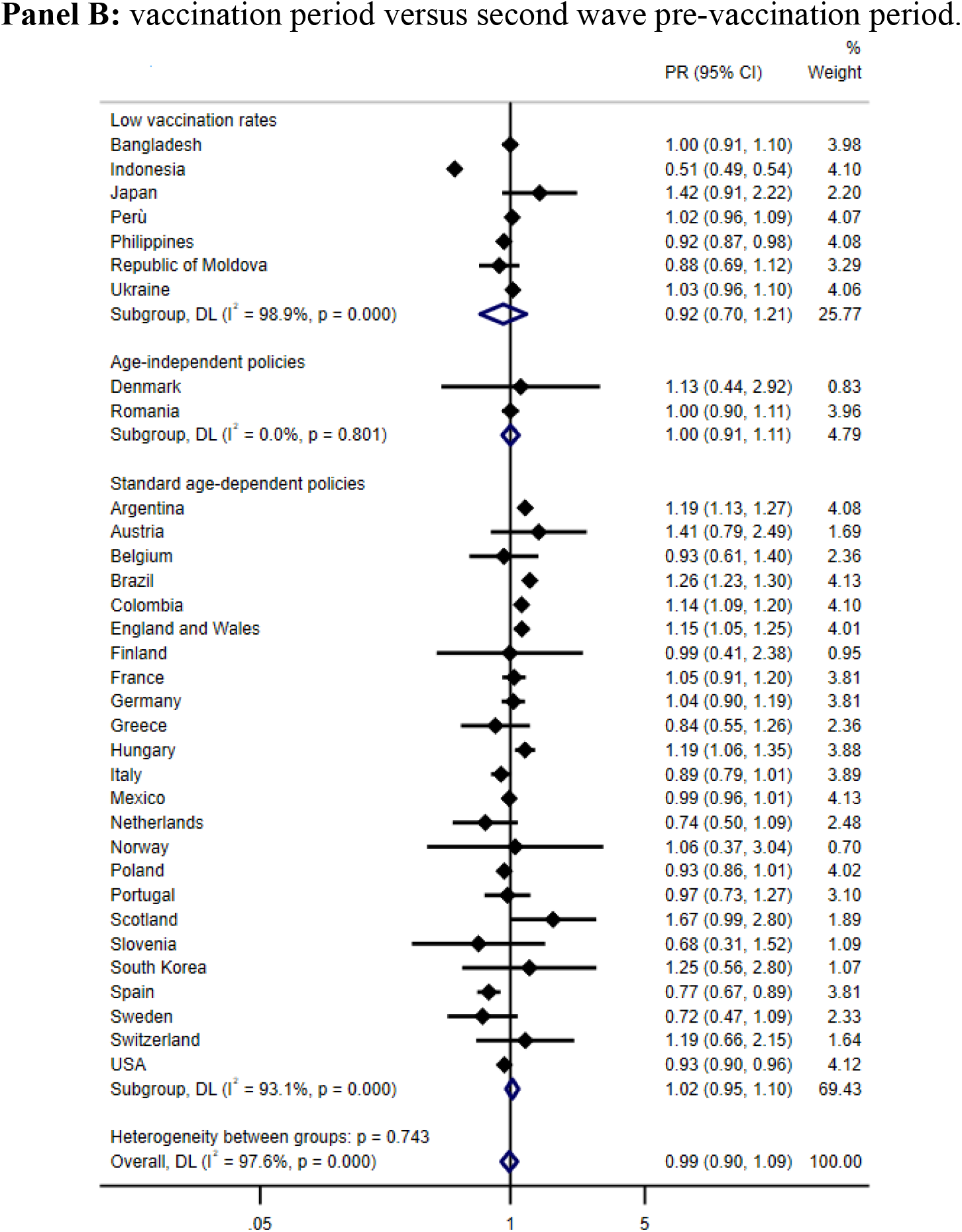
Meta-analysis of prevalence ratios (PR) of COVID-19 deaths in age 0-49 among 0-69 deaths in the vaccination versus pre-vaccination control periods stratified by three main categories 1) Low vaccination rates: countries with a proportion of vaccination with at least one dose < 5% in the general population by the end of April 2021; 2) Age-independent policies: countries with early vaccination policies that vaccinated vulnerable groups regardless of age on 15 January 2021; 3) Standard age-dependent policies: remaining countries. PR >1 implies a higher proportion of deaths accounted by 0-49 age group in the vaccination period versus pre-vaccination periods.

### Regression analyses

The regression analyses provided a suggestive relationship for the prevalence ratio of deaths in the non-elderly in the vaccination versus control pre-vaccination periods and the difference of the proportion of fully vaccinated people in the ≥ 70 versus the 0-69 years group (p=0·018 and p=0·009) (Figure 3). The difference of the proportions of people vaccinated with at least one dose between the two age strata had a significant association with the prevalence ratio of deaths in the non-elderly (p<0·0001 and p<0·0001) (Figure 3).

**Figure 3.**
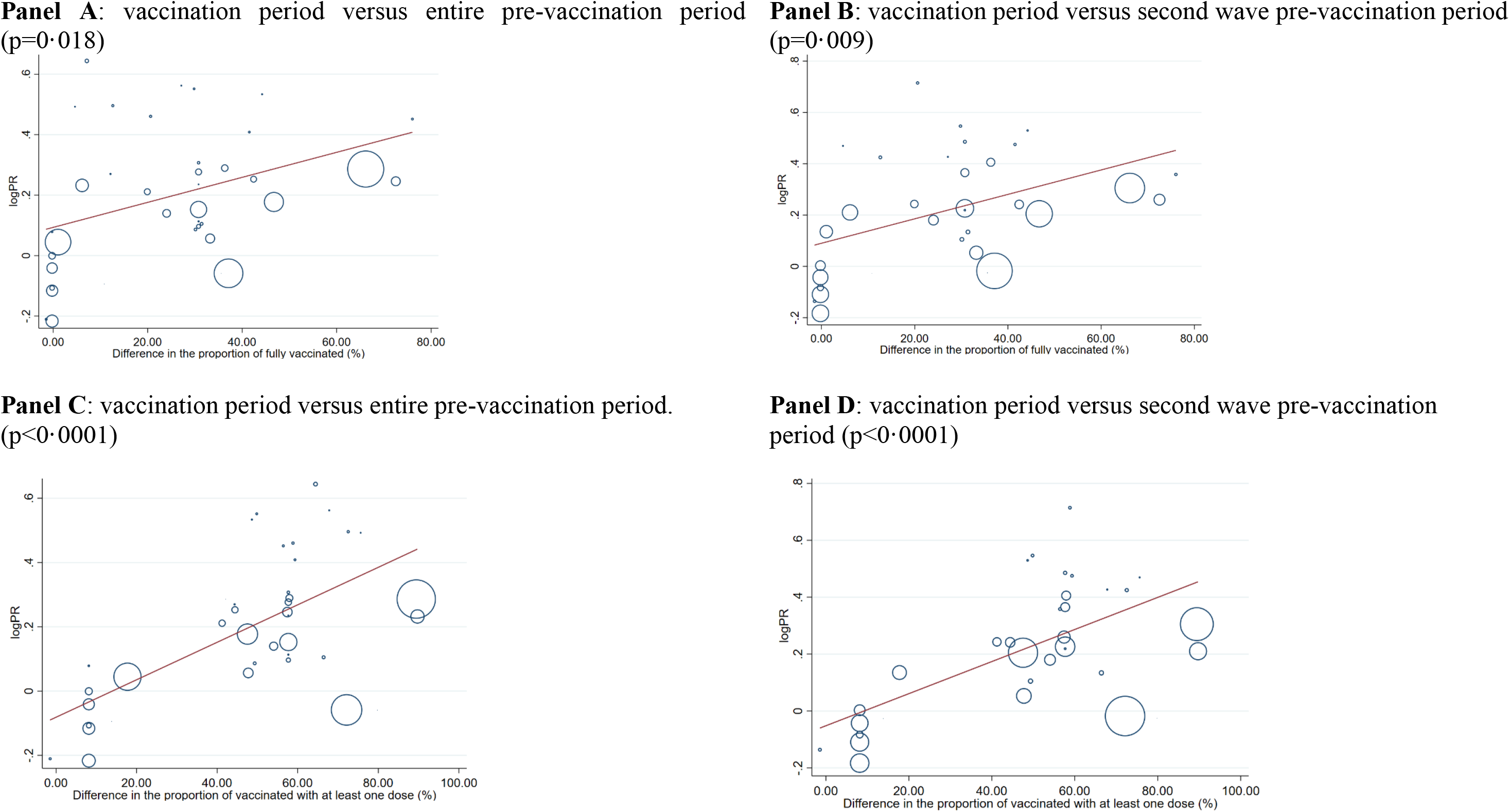
Meta-regression of the logarithm of the prevalence ratio of COVID-19 deaths in the 0-69 age group in the vaccination versus pre-vaccination periods as a function of the difference in the proportion of fully vaccinated people in the ≥ 70 versus 0-69 age strata (Panel A and B) and as a function of the difference in the proportion of vaccinated people with at least one dose in the ≥ 70 versus 0-69 age strata (Panel C and D).

## DISCUSSION

Data from 40 countries show that the use of anti-SARS-CoV-2 vaccination was associated with a marked change in the age distribution of COVID-19 deaths in the first 5 months of 2021 in countries that prioritized vaccination among older people, with a relative increase in the share of deaths among non-elderly people. Conversely, on average, there was no change in the age distribution of COVID-19 deaths during the same time period in countries adopting very early vaccination policies that aimed to vaccinate all the vulnerable people regardless of age and in countries that had very limited use of vaccination.

For countries that prioritized vaccination among elderly people, the increase in the proportion of deaths among the non-elderly was about 1·35-fold. Conversely, there was no change in the relative proportion of COVID-19 deaths among those 0-49 versus those 50-69 years old, which is consistent with the fact that vaccination rates in these two groups did not differ markedly in the first 4 months of 2021 in the vast majority of countries. Exploratory regression suggested that the age shift with a larger share of deaths among the non-elderly was associated with how stark the difference in vaccination rates had been in the elderly versus non-elderly strata. The association was seen most clearly when we considered the difference in the proportion of receiving at least one vaccine dose, with less clear association when data on the rates of full vaccination were examined. This may reflect the fact that a single dose of the widely used vaccines already confers within 2 weeks substantial protection. Moreover, the proportion of people fully vaccinated was much smaller, even in the most elderly strata, probably making the association more difficult to document.

These data offer indirect, but strong support for the ability of vaccination programs to decrease COVID-19 mortality as they can change substantially the death demographics. Apparently, vaccination markedly reduced the population fatality burden among the elderly, the age group carrying the lion’s share of the mortality burden since the beginning of the pandemic. However, it should be noted that despite the substantial age shift in fatalities, the elderly continued to represent the vast majority of deaths in most high-income countries despite the rapid vaccination of a large share of this group. The residual death burden may reflect the fact that many elderly were still not vaccinated in these countries during the period of interest; furthermore, some vulnerable elderly may have launched inefficient immune responses to vaccination (20). Hopefully, the former reason may be the more common one, as clinical trials suggest very good immunogenicity in the elderly for several COVID-19 vaccines. Thorough vaccination of the elderly may dramatically decrease the total footprint of COVID-19 fatalities.

There is no reason to believe that COVID-19 vaccination may not be equally effective (if not more effective) in curtailing fatalities at the population level among non-elderly individuals. During the pandemic, the non-elderly have accounted for a small portion of deaths in high-income countries. Conversely, among the countries that we evaluated, deaths at age <70 have been the majority of documented COVID-19 deaths during the pandemic in Bangladesh, Mexico, Peru, and Moldova, and they have represented 40-50% of the COVID-19 deaths in Brazil, Colombia, Indonesia, Jamaica, and Ukraine. Mortality data for these countries need to be seen with some caution, as documentation of COVID-19 may have been more incomplete. If anything, in several of these countries like Indonesia, Mexico, Philippines, Moldova and Ukraine, there was a suggestion that the proportion of elderly COVID-19 deaths increased in 2021 versus in 2020, despite no or little vaccination of the population. These changes may represent changes in the documentation of COVID-19 deaths, especially among the elderly, e.g. more deaths missed in 2020 due to insufficient testing – or other data artefacts.

Some other limitations need to be discussed. The results presented here represent exploratory analyses of ecological nature and they carry the risk of ecological fallacy and unmeasured, unaccounted confounding. Nevertheless, the patterns observed are compatible and extend the results of large-scale population cohort analyses from countries like Israel and UK that demonstrated large clinical benefits from rapid massive deployment of vaccination in whole countries (4–6). One particular source of unaccounted confounding is the extent to which COVID-19 deaths in nursing home residents may have decreased over time, due to non-vaccine-related reasons, e.g. better protection of nursing homes after the first wave in 2020 and a reduction due to harvesting effects of the population pool of frail nursing home residents who would be highly likely to succumb upon infection. A previous analysis comparing the first versus the second wave (until mid-January 2021) found a large decrease in the proportion of COVID-19 represented by nursing home residents in several high-income countries (9). Available data from several European countries and Canada suggest a further decline in the share of deaths represented by nursing home residents during 2021 (Appendix 1). Universal vaccination is easier to achieve in nursing homes than among the community-dwelling elderly. Nevertheless, deaths seem to have declined equally steeply also among community-dwelling elderly (Appendix 1).

Other potential sources of unaccounted confounding are differences in other measures taken during the vaccination period versus previous periods and the increasing circulation of new variants with different infectiousness and fatality risk (21–24). Non-pharmaceutical interventions changed continuously over time and across countries during the pandemic, and some of these measures may have resulted in shifting deaths to different age strata (25). E.g., relaxation of measures in 2021 may have been more prominent among non-elderly populations. This hypothesis is difficult to prove or disprove, but careful, serial seroprevalence studies may help understand the potential change in population spread of the infection during 2021 versus 2020. Moreover, new variants did circulate widely in many countries during early 2021 (23). In particular, the Gamma variant became widely spread in South America (26) and the Alpha variant became widespread in many high-income countries (24). There is some tentative evidence for increased infectivity (27) and even potentially increased mortality risk (28). It is also possible that infection by some variants may be less likely to be prevented by prior infection (29). However, it is unclear whether these features would have affected the mortality footprint more prominently among younger populations.

Finally, not all COVID-19 vaccines may be equally effective in curbing fatalities and effectiveness for decreasing mortality of some vaccines may vary across different age strata. Of note, a most prominent outlier in our analyses was Mexico, where the proportion of elderly COVID-19 deaths increased during 2021. Mexico has used 6 different vaccines, including Chinese vaccines (Sinovac/Sinopharm) that may have lower effectiveness than other licensed vaccines (30).

Acknowledging these caveats, our analyses show a substantial shift in the age distribution of COVID-19 deaths in countries that rapidly implemented vaccination predominantly among their elderly populations. Lower total fatalities and a shift towards the younger population probably represent favorable news of population-level vaccination effectiveness and a transition of the pandemic towards less threatening endemicity. Nevertheless, the demographic features of COVID-19 fatalities need to continue to be carefully scrutinized over time.

## Data Availability

All data used in this work are shown in the tables and supplementary materials and were collected from national and international databases.

## RESEARCH IN CONTEXT

### Evidence before this study

Several COVID-19 vaccines have shown high efficacy and safety in large clinical trials and therefore have been authorized for wide use in the general population. The global COVID-19 vaccination campaign has differed nationally, varying in timing and setting of priority categories. We searched PubMed using the terms “COVID-19”, “deaths”, “age distribution” or “demographics”, and respective synonyms without language or date restrictions. We identified national policies on vaccination deployment and COVID-19 death reporting searching targeted websites and consulting with national experts. The proportion of COVID-19 deaths in the younger population was higher in the low- and middle-income countries than in high-income countries. Age distribution of COVID-19 deaths was similar throughout 2020 in the so-called “first wave” and “second wave” in high-income countries, but it has been insufficiently studied in low- and middle-income countries, and during the period after the first vaccines were used in the general population.

### Added value of this study

This study is the most comprehensive analysis to date of global COVID-19 deaths age distribution integrating data for deaths and vaccination from diverse publicly available national and international sources. Our study points to population-level effectiveness of COVID-19 vaccination and shows that vaccination policies can change substantially COVID-19 deaths demographics. Non-protected population groups have a larger share of COVID-19 deaths, as deaths decrease in vaccine-protected strata.

### Implications of all the available evidence

While the demographic features of COVID-19 fatalities need to continue to be scrutinized as the pandemic evolves, our results inform citizens, policymakers, and researchers worldwide about the potential effect of COVID-19 vaccination policies and a favorable evolution of the pandemic towards endemicity with a much lower total fatality burden.

---

**Table S1.** Sources about COVID-19 deaths and date of trough, separation date and last date available.

| Country | Date of trough | Separation date | Latest date | Source |
| --- | --- | --- | --- | --- |
| Argentina | Oct 1 2020 | Oct 1 2020 | June 2 2021 | Riffe T, Acosta E. COVerAGE-DB: A database of COVID-19 cases and deaths by age. doi: 10-17605/OSF. IO/MPWJQ. 2020. |
| Australia | May 9 2020 | May 9 2020 | June 4 2021 | Riffe T, Acosta E. COVerAGE-DB: A database of COVID-19 cases and deaths by age. doi: 10-17605/OSF. IO/MPWJQ. 2020. |
| Austria | June 5 2020 | June 5 2020 | May 27 2021 | Institut national d'etudes demographiques (INED). available at: https://dc-covid.site.ined.fr/en/data/ |
| Bangladesh | May 6 2020 | June 2 2020 | Apr 29 2021 | Riffe T, Acosta E. COVerAGE-DB: A database of COVID-19 cases and deaths by age. doi: 10-17605/OSF. IO/MPWJQ. 2020. |
| Belgium | July 14 2020 | July 14 2020 | May 26 2021 | Institut national d'etudes demographiques (INED). available at: https://dc-covid.site.ined.fr/en/data/ |
| Brazil | Nov 11 2020 | Nov 11 2020 | Apr 19 2021 | Riffe T, Acosta E. COVerAGE-DB: A database of COVID-19 cases and deaths by age. doi: 10-17605/OSF. IO/MPWJQ. 2020. |
| Canada | Sept 7 2020 | NA | May 21 2021 | Institut national d'etudes demographiques (INED). available at: https://dc-covid.site.ined.fr/en/data/ |
| Colombia | Oct 10 2020 | Oct 10 2020 | June 2 2021 | Riffe T, Acosta E. COVerAGE-DB: A database of COVID-19 cases and deaths by age. doi: 10-17605/OSF. IO/MPWJQ. 2020. |
| Denmark | June 17 2020 | June 17 2020 | May 27 2021 | Institut national d'etudes demographiques (INED). available at: https://dc-covid.site.ined.fr/en/data/ |
| England and Wales | Aug 21 2020 | Aug 21 2020 | May 14 2021 | Institut national d'etudes demographiques (INED). available at: https://dc-covid.site.ined.fr/en/data/ |
| Finland | June 6 2020 | June 6 2020 | June 2 2021 | Riffe T, Acosta E. COVerAGE-DB: A database of COVID-19 cases and deaths by age. doi: 10-17605/OSF. IO/MPWJQ. 2020. |
| France | Aug 10 2020 | Aug 10 2020 | May 27 2021 | Institut national d'etudes demographiques (INED). available at: https://dc-covid.site.ined.fr/en/data/ |
| Germany | Aug 1 2020 | Aug 1 2020 | May 25 2021 | Institut national d'etudes demographiques (INED). available at: https://dc-covid.site.ined.fr/en/data/ |
| Greece | May 30 2020 | May 30 2020 | May 26 2021 | Riffe T, Acosta E. COVerAGE-DB: A database of COVID-19 cases and deaths by age. doi: 10-17605/OSF. IO/MPWJQ. 2020. |
| Honduras | Dec 3 2020 | Dec 3 2020 | Apr 21 2021 | Riffe T, Acosta E. COVerAGE-DB: A database of COVID-19 cases and deaths by age. doi: 10-17605/OSF. IO/MPWJQ. 2020. |
| Hungary | July 7 2020 | July 7 2020 | June 2 2021 | Riffe T, Acosta E. COVerAGE-DB: A database of COVID-19 cases and deaths by age. doi: 10-17605/OSF. IO/MPWJQ. 2020. |
| Indonesia | Aug 15 2020 | Aug 15 2020 | June 4 2021 | Riffe T, Acosta E. COVerAGE-DB: A database of COVID-19 cases and deaths by age. doi: 10-17605/OSF. IO/MPWJQ. 2020. |
| Israel | May 27 2020 | May 27 2020 | Apr 18 2021 | Riffe T, Acosta E. COVerAGE-DB: A database of COVID-19 cases and deaths by age. doi: 10-17605/OSF. IO/MPWJQ. 2020. |
| Italy | Aug 15 2020 | Aug 18 2020 | May 26 2021 | Institut national d'etudes demographiques (INED). available at: https://dc-covid.site.ined.fr/en/data/ |
| Jamaica | NA | NA | June 2 2021 | Riffe T, Acosta E. COVerAGE-DB: A database of COVID-19 cases and deaths by age. doi: 10-17605/OSF. IO/MPWJQ. 2020. |
| Japan | July 6 2020 | July 6 2020 | May 24 2021 | National Institute of Population and Social Security Research. Available at: http://www.ipss.go.jp/index-e.asp |
| Jordan | NA | NA | June 4 2021 | Riffe T, Acosta E. COVerAGE-DB: A database of COVID-19 cases and deaths by age. doi: 10-17605/OSF. IO/MPWJQ. 2020. |
| Mexico | Oct 10 2020 | Oct 10 2020 | May 3 2021 | Riffe T, Acosta E. COVerAGE-DB: A database of COVID-19 cases and deaths by age. doi: 10-17605/OSF. IO/MPWJQ. 2020. |
| Netherlands | July 14 2020 | July 14 2020 | May 25 2021 | Institut national d'etudes demographiques (INED). available at: https://dc-covid.site.ined.fr/en/data/ |
| Northern Ireland | NA | NA | June 3 2021 | Riffe T, Acosta E. COVerAGE-DB: A database of COVID-19 cases and deaths by age. doi: 10-17605/OSF. IO/MPWJQ. 2020. |
| Norway | May 21 2020 | May 22 2020 | May 27 2021 | Institut national d'etudes demographiques (INED). available at: https://dc-covid.site.ined.fr/en/data/ |
| Peru | Dec 3 2020 | Dec 3 2020 | June 2 2021 | Riffe T, Acosta E. COVerAGE-DB: A database of COVID-19 cases and deaths by age. doi: 10-17605/OSF. IO/MPWJQ. 2020. |
| Philippines | May 24 2020 | May 24 2020 | June 1 2021 | Riffe T, Acosta E. COVerAGE-DB: A database of COVID-19 cases and deaths by age. doi: 10-17605/OSF. IO/MPWJQ. 2020. |
| Poland | July 22 2020 | July 22 2020 | May 24 2021 | Mapy potrzeb zdrowotnych - Ministerstwo Zdrowia. Available at: https://basiw.mz.gov.pl/index.html#/visualization?id=3653 |
| Portugal | Aug 7 2020 | Aug 7 2020 | May 26 2021 | Institut national d'etudes demographiques (INED). available at: https://dc-covid.site.ined.fr/en/data/ |
| Republic of Moldova | Aug 24 2020 | Aug 24 2020 | May 27 2021 | Institut national d'etudes demographiques (INED). available at: https://dc-covid.site.ined.fr/en/data/ |
| Romania | June 3 2020 | June 3 2020 | May 27 2021 | Institut national d'etudes demographiques (INED). available at: https://dc-covid.site.ined.fr/en/data/ |
| Scotland | Aug 30 2020 | Aug 30 2020 | May 23 2021 | Institut national d'etudes demographiques (INED). available at: https://dc-covid.site.ined.fr/en/data/ |
| Slovenia | May 10 2020 | May 10 2020 | May 30 2021 | Nacionalni inštitut za javno zdravje. Available at: https://www.nijz.si/sl/dnevno-spremljanje-okuzb-s-sars-cov-2-covid-19 |
| South Korea | May 12 2020 | Apr 15 2020 | May 26 2021 | Institut national d'etudes demographiques (INED). available at: https://dc-covid.site.ined.fr/en/data/ |
| Spain | July 27 2020 | July 27 2020 | May 26 2021 | Institut national d'etudes demographiques (INED). available at: https://dc-covid.site.ined.fr/en/data/ |
| Sweden | Aug 28 2020 | Aug 31 2020 | May 23 2021 | Institut national d'etudes demographiques (INED). available at: https://dc-covid.site.ined.fr/en/data/ |
| Switzerland | June 1 2020 | June 2 2020 | May 27 2021 | Institut national d'etudes demographiques (INED). available at: https://dc-covid.site.ined.fr/en/data/ |
| Ukraine | June 3 2020 | June 3 2020 | May 27 2021 | Institut national d'etudes demographiques (INED). available at: https://dc-covid.site.ined.fr/en/data/ |
| Uruguay | May 25 2020 | May 25 2020 | May 27 2021 | Github. Available at: https://github.com/butaca/covid19uy.com |
| USA | July 5 2020 | July 4 2020 | May 26 2021 | Institut national d'etudes demographiques (INED). available at: https://dc-covid.site.ined.fr/en/data/ |

**Table S2.**
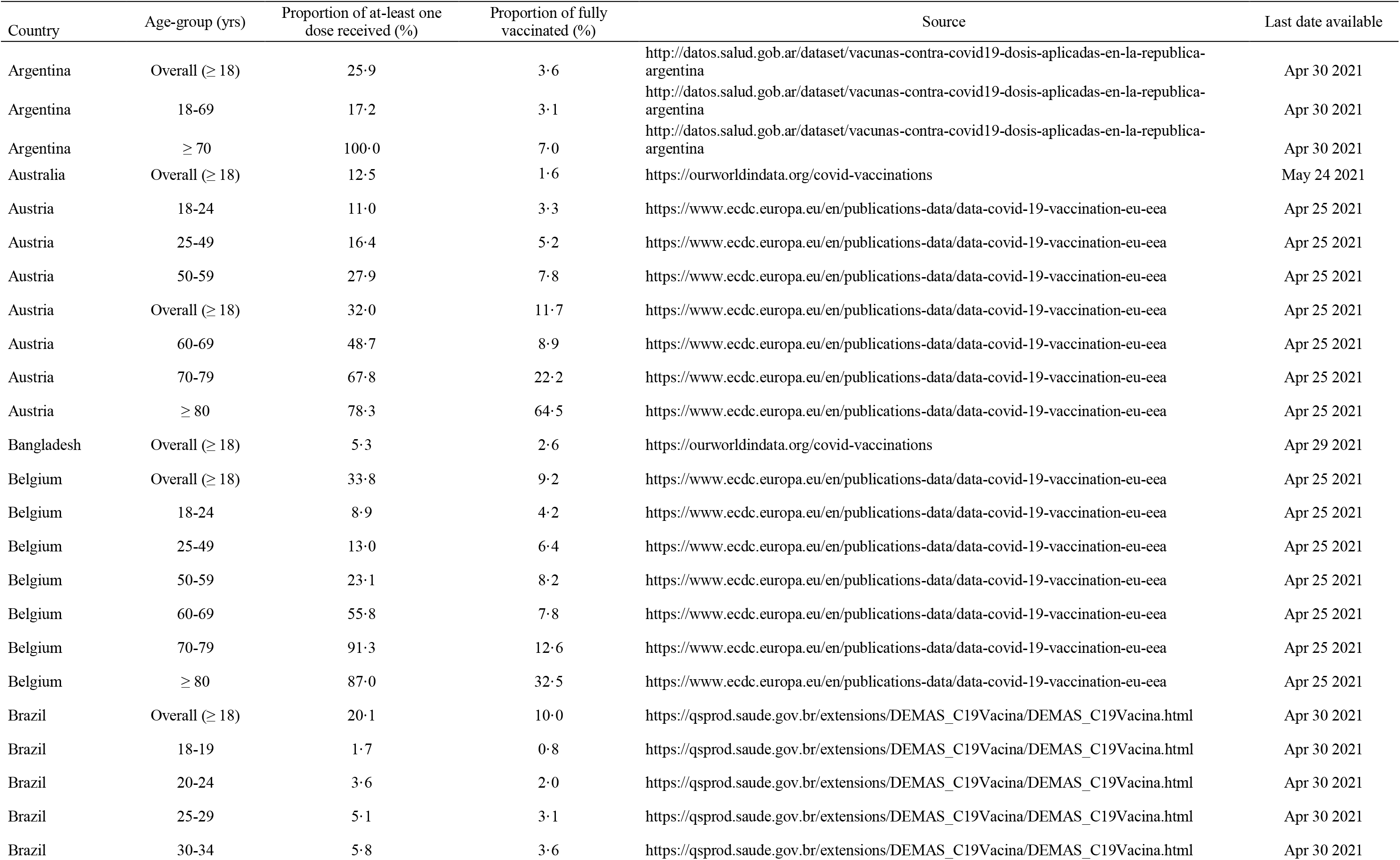
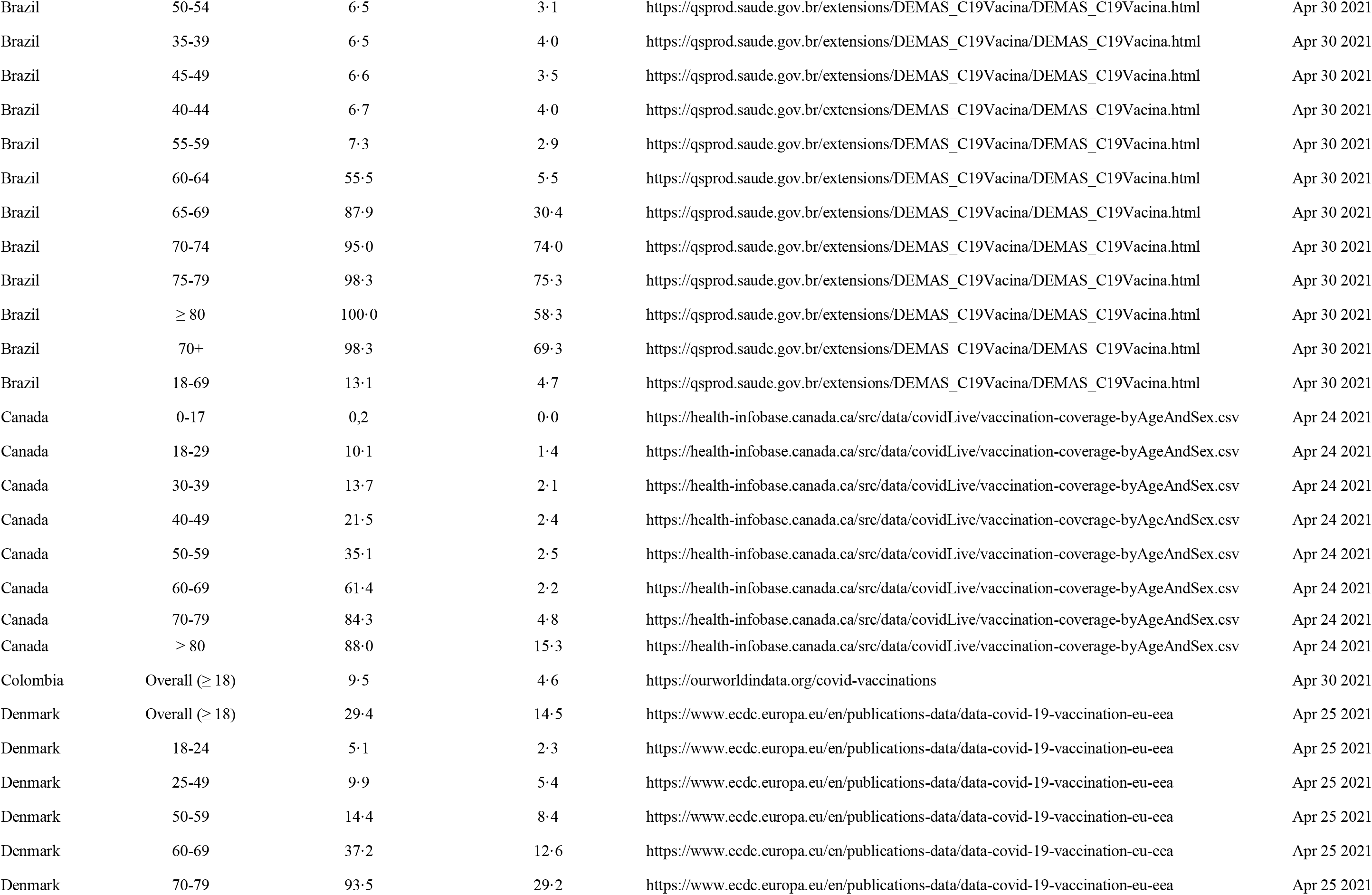
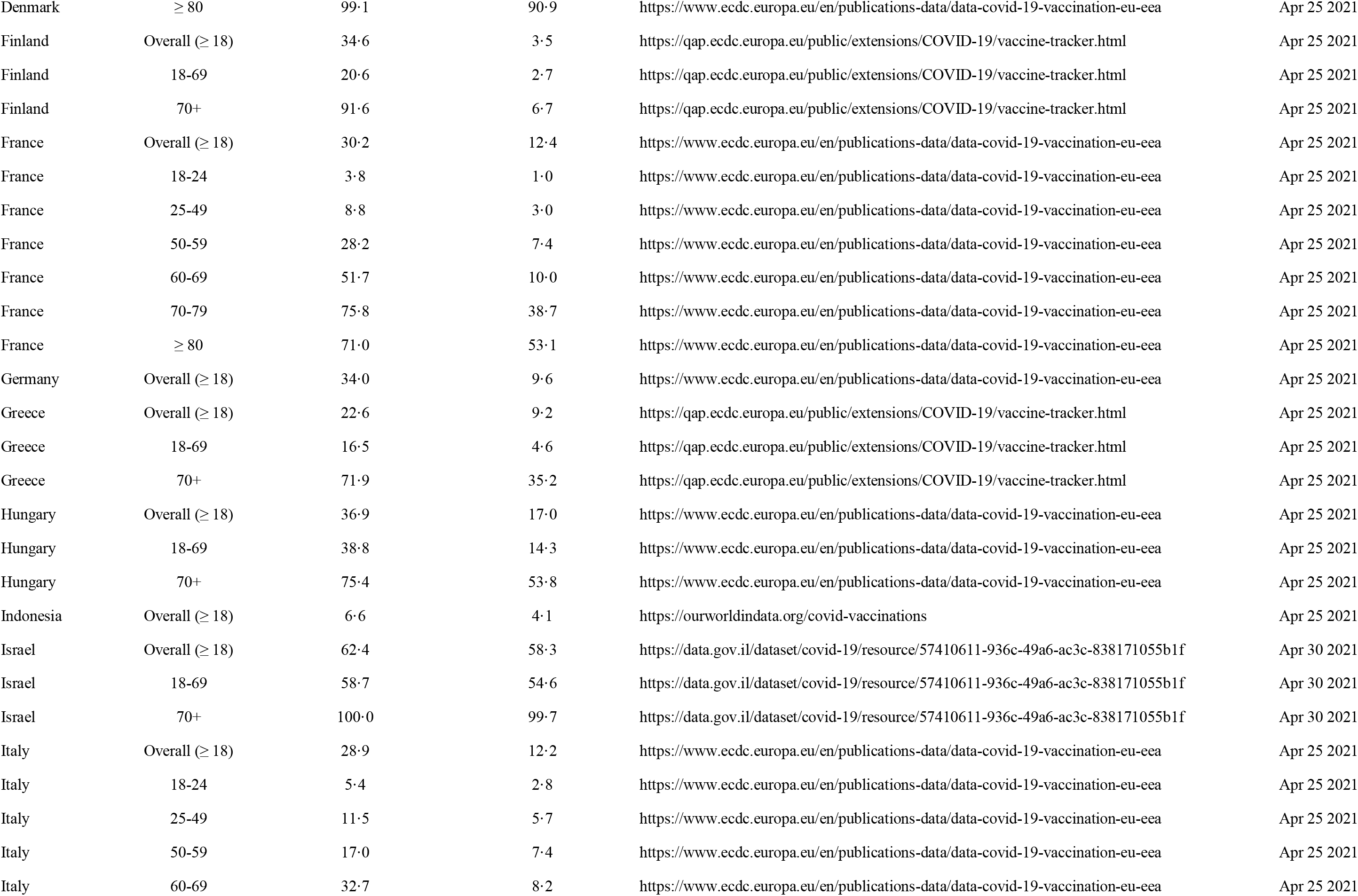
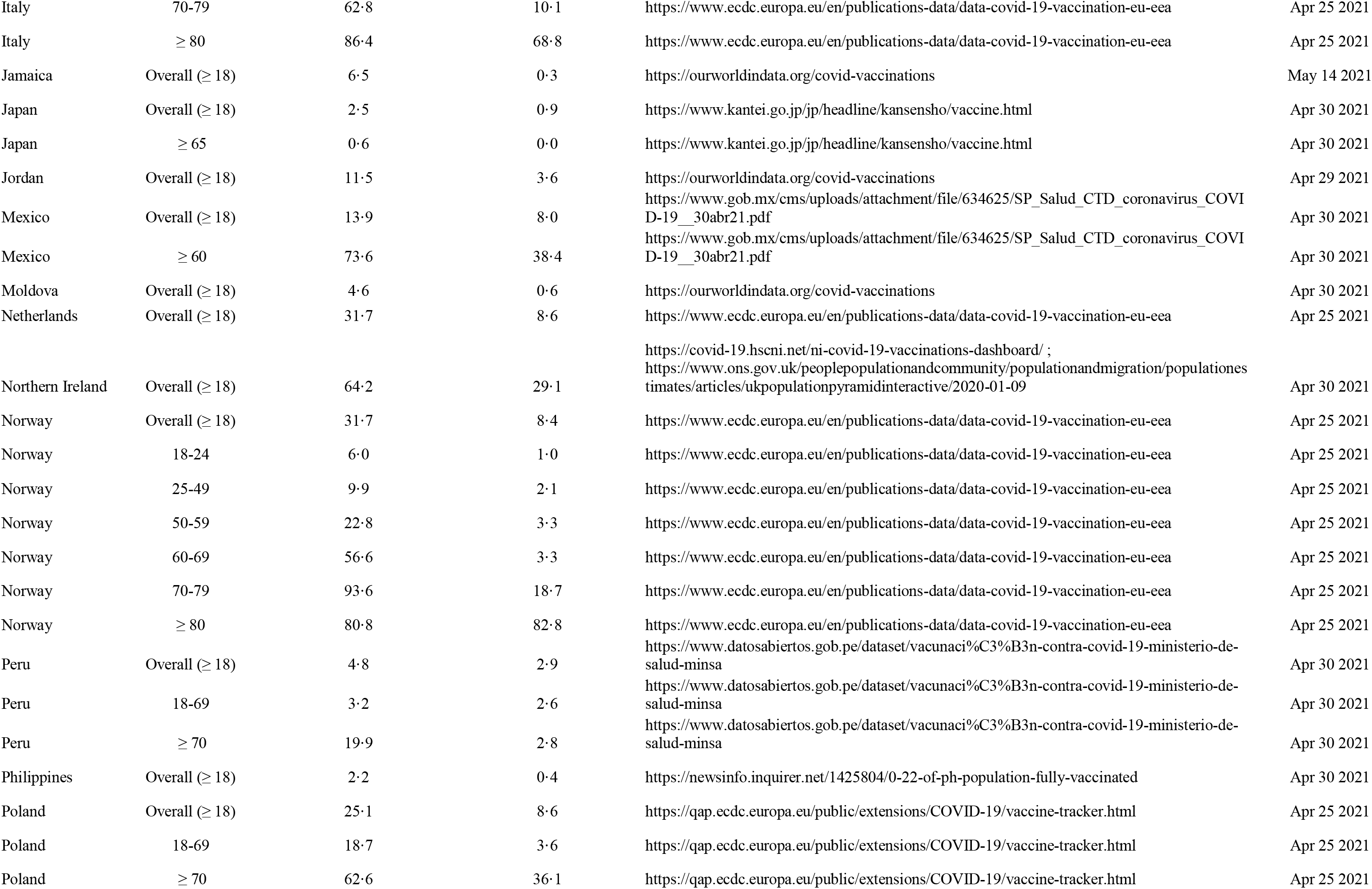
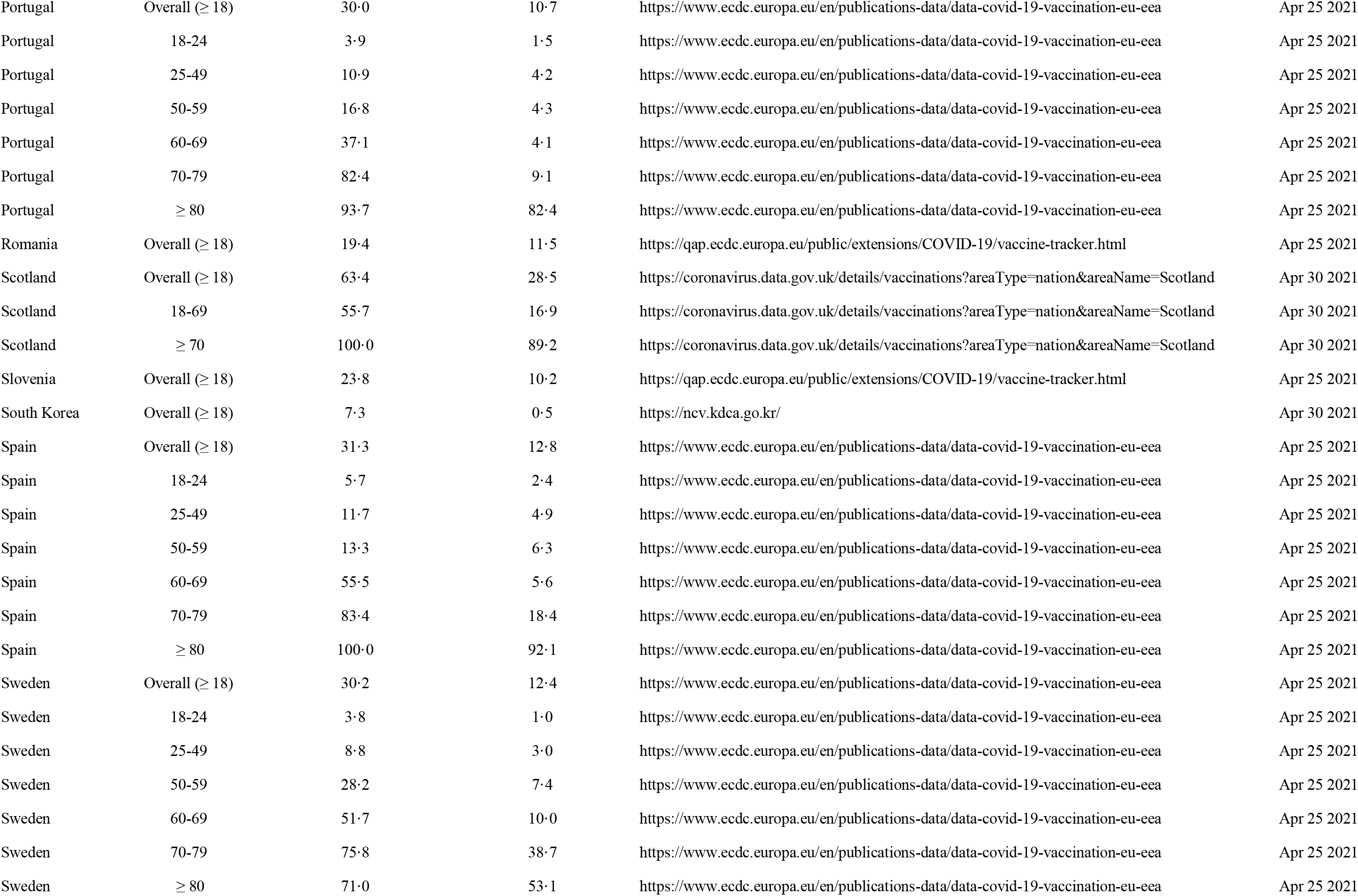
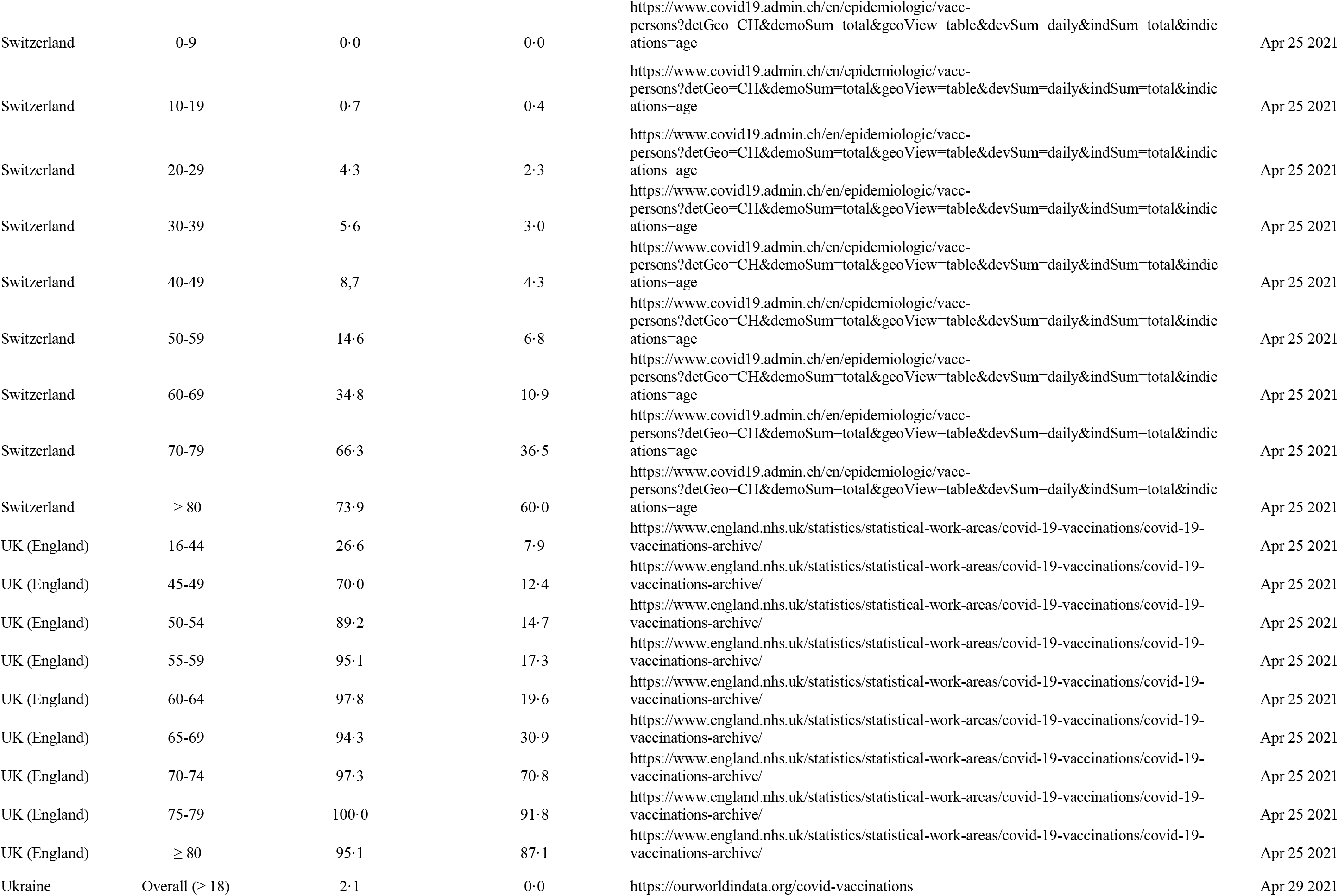
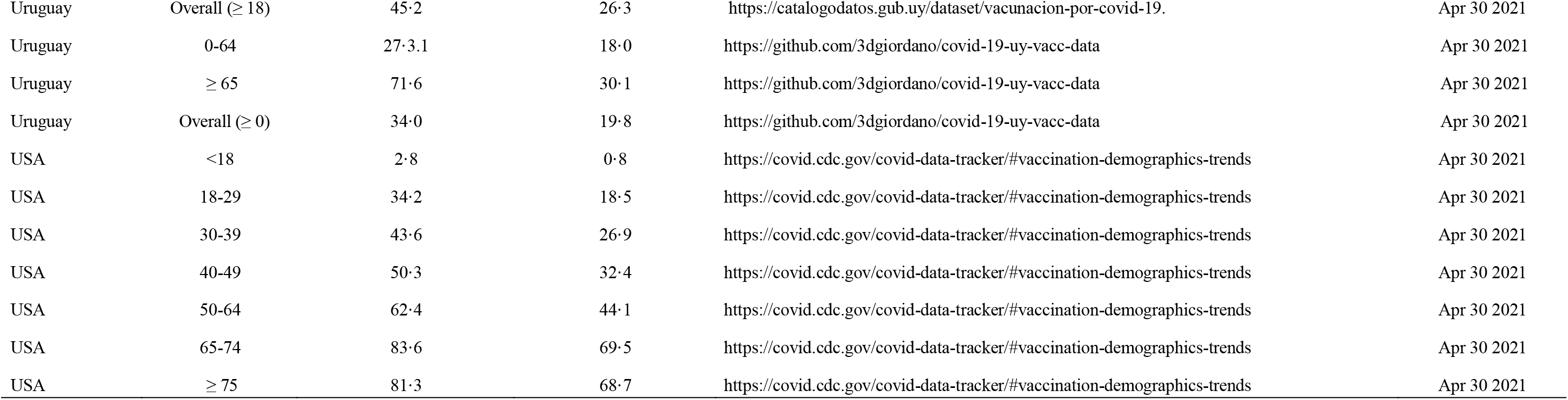
COVID-19 vaccination coverages, sources and last date available

**Table S3.**
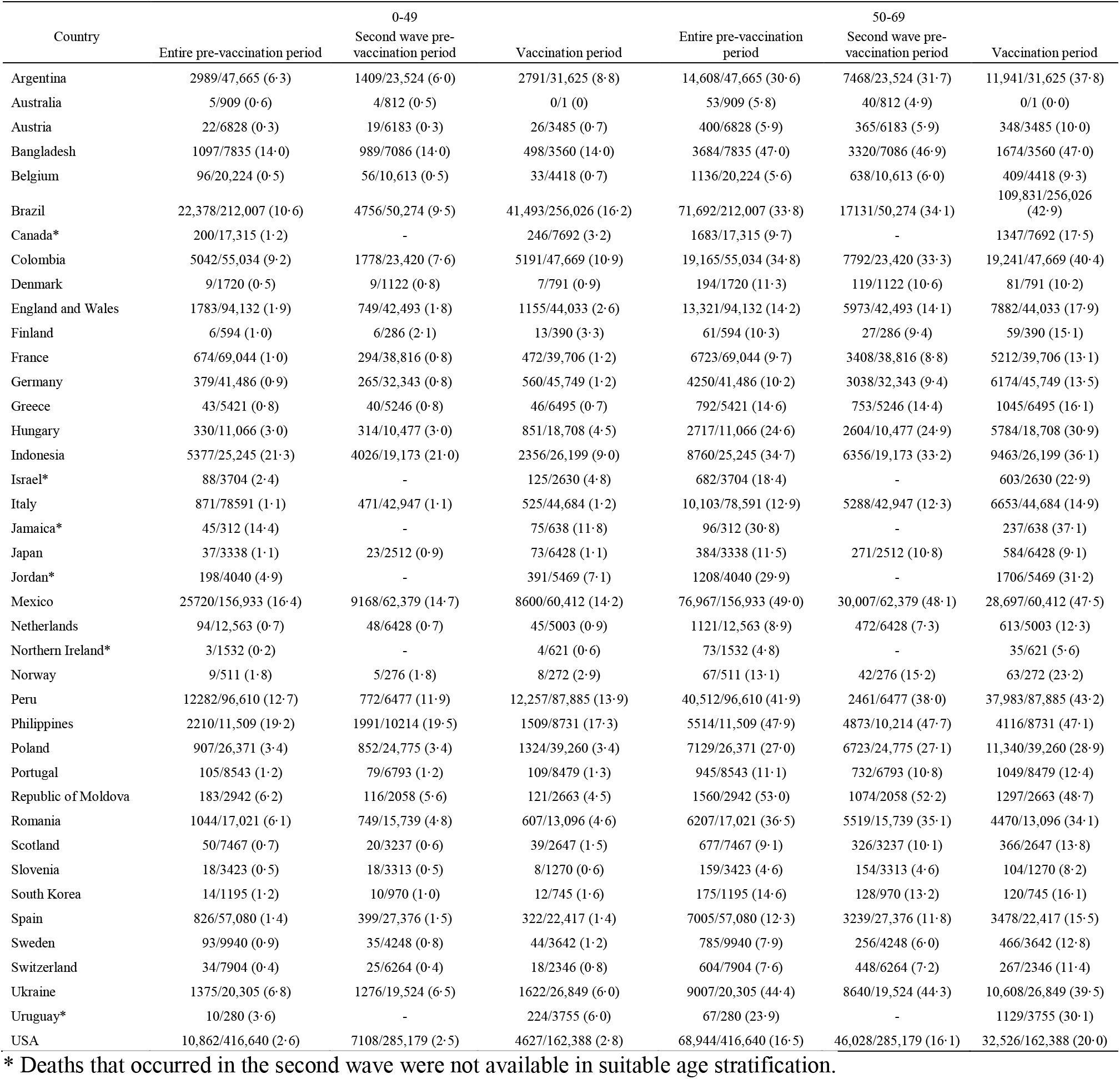
Proportion of COVID-19 deaths in the 0-49 and 50-69 age groups to total COVID-19 deaths.

## Declarations

### Author Contributions

All authors contributed to the study conception and design. Material preparation and data collection were performed by Roberta Pastorino, Leonardo Villani, Angelo Maria Pezzullo, Francesco Andrea Causio, Cathrine Axfors, Despina G. Contopoulos-Ioannidis. Roberta Pastorino and Angelo Maria Pezzullo performed the statistical analysis. The first draft of the manuscript was written by John P.A. Ioannidis, Despina G. Contopoulos-Ioannidis, Leonardo Villani, Roberta Pastorino, Angelo Maria Pezzullo and Stefania Boccia. John P.A. Ioannidis, Cathrine Axfors, Despina G. Contopoulos-Ioannidis and Stefania Boccia commented on the subsequent and latest version of the manuscript. John P.A. Ioannidis supervised the work. All authors have read and agreed to the published version of the manuscript.

## Acknowledgements

We would like to thank Juan Fornos for kindly providing the Uruguay data he extracted from official sources.

## Ethics approval

Not applicable.

## APPENDIX 1

We considered the proportion of nursing home COVID-19 deaths in eleven countries (Australia, Belgium, Canada, Denmark, Finland, France, Germany, Norway, Sweden, UK, USA) in the first wave and second wave from a previous analysis (1) and added the proportion of deaths for the vaccination period (from the end date of second wave to May 31 or more proximal date) using the same sources for consistency purposes. We could find updated data for ten countries (all countries except Finland). We did not considered Australia because it has not registered any COVID-19 death in nursing homes the vaccination period. When only the number of nursing home COVID-19 deaths was available, we obtained the total number of COVID-19 deaths in the same period from Our World in Data database (2) to obtain the proportion (Appendix Table 1).

**Appendix Table 1.** Proportion of COVID-19 deaths occurring in nursing home residents.

| Country | Proportion of nursing home resident deaths in FW (%) | End date of FW (2020) | Proportion of nursing home resident deaths in SW (%) | End date of SW (2021) | Proportion of nursing home resident deaths in VP (%) | Latest date (2021) | Definition of nursing home deaths \$ | Definition of nursing home and other included facilities |
| --- | --- | --- | --- | --- | --- | --- | --- | --- |
| Belgium | 50.5 | June 20 | 36.8 | Jan 10 | 15.4 | May 30 | O, C/P | Nursing homes |
| Canada | 85.1 | June 1 | 59.0 | Jan 14 | 38.7 | Jul 7 | R, C/P | Nursing and retirement homes |
| Denmark | 35.3 | June 15 | 39.0 | Jan 12 | 37.3 | Jul 5 | R, C | Nursing homes |
| France | 48.5 | June 16 | 17.4 | Jan 10 | 13.6 | May 23 | R, C/P | Social and medico-social establishments (ESMS) |
| Germany | 39.2 | June 23 | 24.3 | Jan 14 | 22.9 | May 25 | R, C | Communal settings (e.g. nursing homes, homeless shelters, accommodation for refugees, prisons) |
| Norway | 59.0 | June 19 | 61.5 | Jan 20 | 36.0 | Jun 2 | O, C | Other health institutions than hospitals: nursing homes and other institutions |
| Sweden | 47.4 | June 15 | 37.1 | Jan 18 | 23.7 | Jul 4 | R, C/P | Nursing homes £ |
| UK | 31.1 | June 14 ¶ | 20.0 | Jan 1§ | 24.2 | May 14 | O, C/P | Nursing homes |
| USA | 45.0 | June 18 | 34.7 | Jan 5 | 25.7 | Jul 2 | R, C/P | Nursing facilities, assisted living facilities, adult care centers, intermediate care centers, and/or long-term care facilities |

Abbreviations: FW, first wave; SW, second wave; VP, vaccination period. # Total number of deaths with information on nursing home residence status or place of death. \$ By place of residence (R) or by place of occurrence (O; i.e., not counting in-hospital deaths of nursing home residents); confirmed COVID-19 (C); confirmed or probable COVID-19 (C/P). ¶ England, Wales, and Northern Ireland: June 12, 2020; Scotland: June 14, 2020. § England and Wales and Northern Ireland: January 1, 2021; Scotland: January 4, 2021. £ Different definitions of place of residence are used for Swedish FW and SW data (change reported by the Swedish National Board of Health and Welfare on August 24, 2020).

We calculated prevalence ratios (PRs) with 95% confidence intervals (CI) for the proportion of nursing home residents COVID-19 deaths among all COVID-19 deaths in the second wave (also called “Period B” in the main paper) for all countries using the COVID-19 death data from the main paper.

**Appendix Figure 1.**
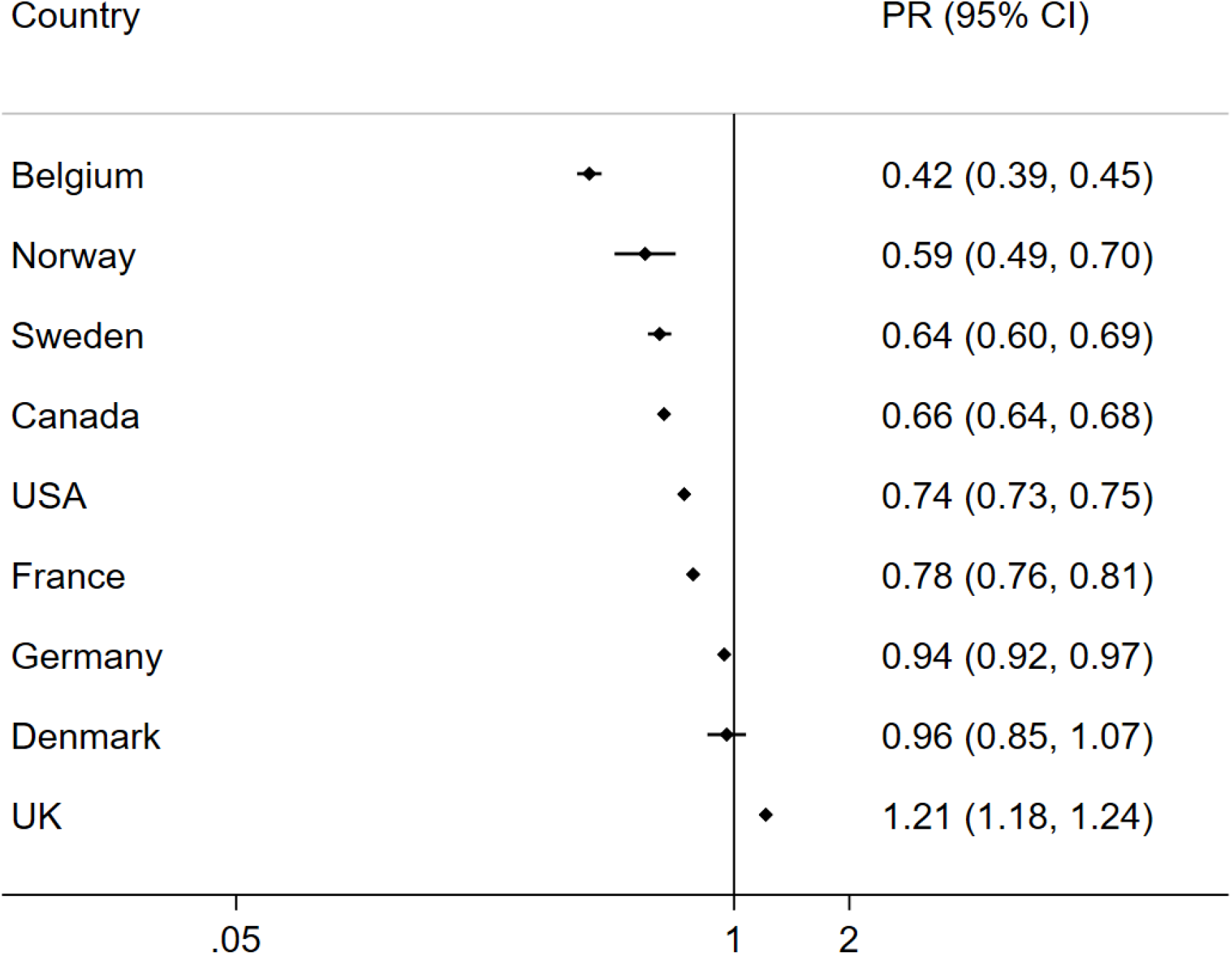
COVID-19 deaths in nursing home residents: prevalence ratio for the second wave versus the vaccination period. PR<1 indicates a decrease in the share of nursing home COVID-19 deaths among all COVID-19 deaths.

The proportion of nursing home COVID-19 deaths among all COVID-19 deaths was lower in the vaccination period than in the second wave in 8 of the 9 countries (Appendix Figure 1). There were statistically significant reductions in 7 countries (prevalence ratios 0.42 to 0.94). There was no significant difference in Denmark, and an increase in the proportion of nursing home COVID-19 deaths in the UK (prevalence ratio 1.21, 95% CI 1.18–1.24) in the vaccination period.

We used the proportion of nursing home deaths in the three periods (first wave, second wave, vaccination period) to estimate the number of age stratified community-dwelling (non-nursing home) deaths in the three periods. To calculate it we subtracted the proportion of nursing home deaths from the total deaths of the main paper, a 5% of the nursing home deaths from the 0-69 age stratum, and a 95% of the nursing home deaths from the ≥d 70 age stratum (Appendix Table 2).

**Appendix Table 2.**
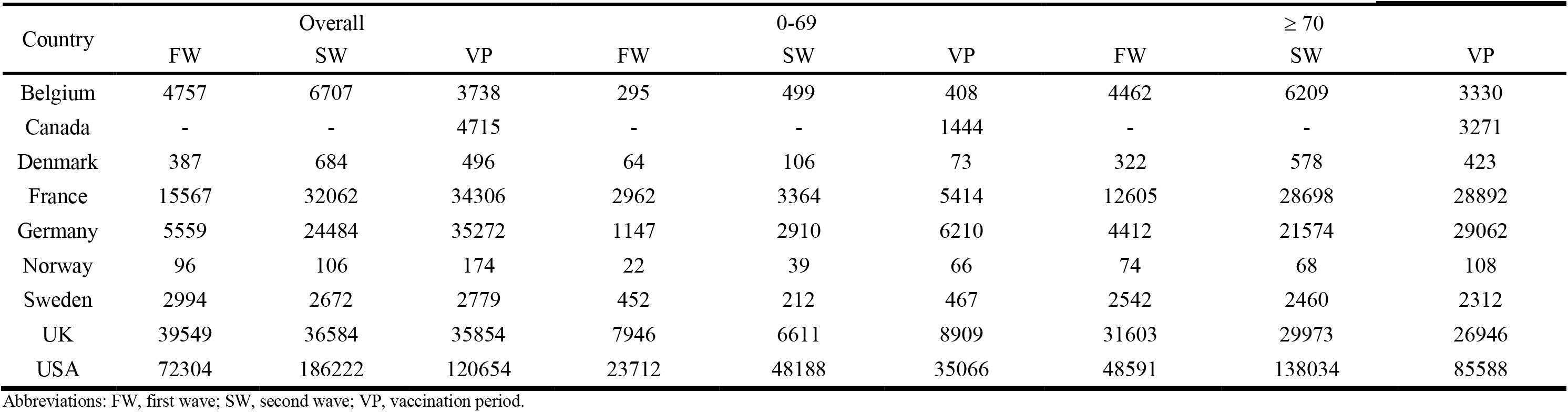
Estimated community-dwelling COVID-19 deaths in the first wave, second wave, and vaccination period

As we did in the main paper, we calculated PRs for the second wave versus the vaccination period for the proportion of COVID-19 deaths in the 0-69 age group among all COVID-19 deaths, using the estimate community-dwelling deaths. The rationale of doing so was to examine if the shift in the demographics of COVID-19 deaths in these countries between the two periods was still present excluding nursing home deaths. In eight countries with available data, six countries had a significant shift in the demographics of COVID-19 deaths when excluding nursing home deaths (Appendix Figure 2, Panel A). We did not observe a shift in the demographics of community deaths in Denmark (consistently with what we found in the general population) and Norway, where few deaths were observed overall. We report a subset of the main analysis (reported in Figure 1 Panel B of the main paper, data for nursing homes are not excluded) for the same eight countries (Appendix Figure 2, Panel B). The summary PRs of the two analyses (estimated community-dwelling deaths versus all deaths) are similar.

**Appendix Figure 2.**
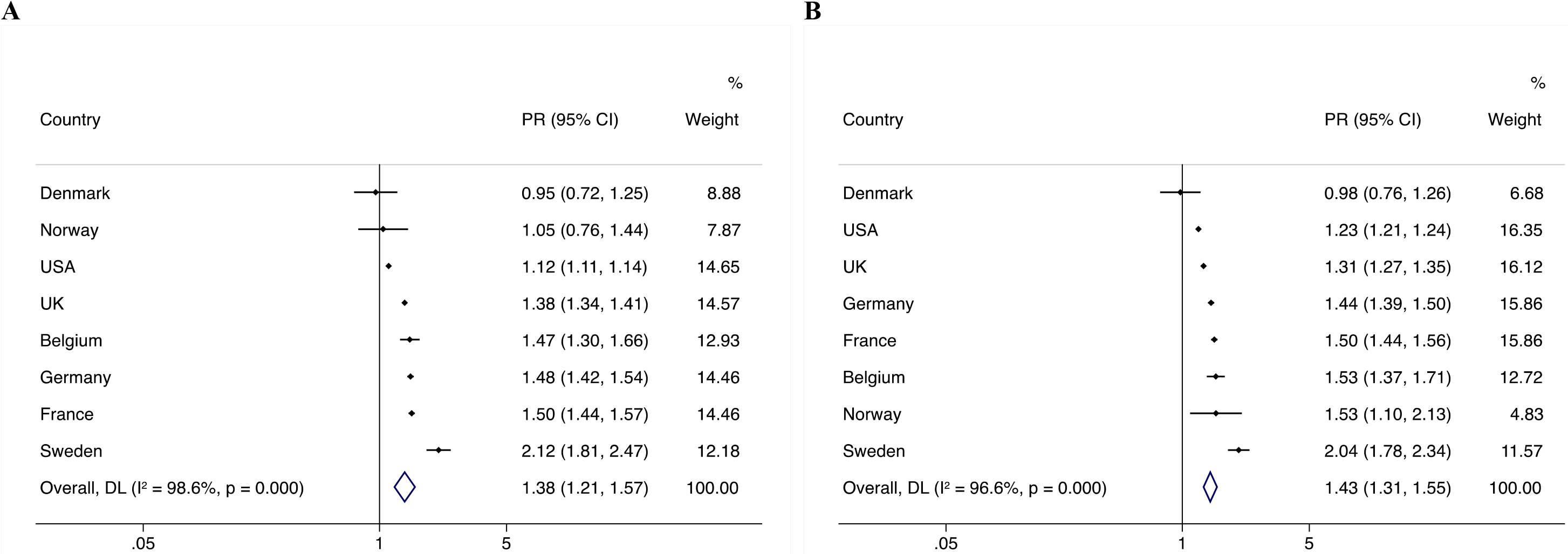
Meta-analysis of prevalence ratios (PR) of 0-69 age group among all deaths for the second wave versus the vaccination period. Panel A, estimated community-dwelling COVID-19 deaths; Panel B, all observed COVID-19 deaths. PR>1 indicates an increase in the share of 0-69 COVID-19 deaths among all COVID-19 deaths.

